# Model Based Comparison of Covid-19 Cases in Two Counties in the Silicon Valley

**DOI:** 10.1101/2020.05.26.20114116

**Authors:** Santanu Basu

## Abstract

A recently developed model to analyze Covid-19 case data has been applied to compare the Covid-19 cases and hospital bed usage in Santa Clara county and San Mateo county which are in the Silicon Valley region of California. The model gives prediction for number of cases and deaths and number of hospital beds six weeks in advance. The model is versatile and can be applied to other countries and regions as well.

## INTRODUCTION

Covid-19 has spread rapidly through all counties in California, USA since the first reported case on January 26^th^, 2020. A mathematical model has been recently developed to analyze the Covid-19 case data and then to predict the number of cases six weeks in advance [1,2]. This model has been applied to many countries and to the state of California. Since treatment of patients is local, it is valuable to know at the county level the expected number of patients who would require hospitalization in the near future. This would enable distributing case load among hospitals and to adequately equip the hospitals with staffing, PPE and life support equipment. Sometimes two neighboring counties may have different outcomes from the same disease. Modelling can provide insight into the origin of the differences. This is the objective of this paper.

## OVERVIEW OF THE MATHEMATICAL MODEL

We developed and documented a mathematical model to predict number of cases and hospital patient loads [1,2]. The model assumes that starting from a small number of infected people which act a seed, the infection grows by transmission through airborne and physical contact means to the general population. The model keeps track of the number of infected people at various stages of development of the disease through the course of the virus outbreak. The model is parametric and transparent so that predictions are based on parameters which are in principle measurable and the predictions can be continuously corrected as more accurate data on parameters become available. Typical machine learning and curve fitting models which are not based on any real-life parameters tend to have high degree of error in making projections and do not provide physical insight into the problem that can be used in the future to minimize casualties. At present, the model has twenty one user-supplied variable parameters, three of which influence the results greatly. These three are (1) the first date of implementation of social distancing protocols since the viral outbreak, (2) the rate at which the rate of transmission decreases by using the government measures and (3) the final rate of transmission in the long run.

In this paper, we will be using the standard model as described above. Work is in progress to include population and patient movement across the county boundaries. The objective of this paper is to analyze the cases of two adjacent counties in California and to predict six weeks in advance the hospitalization requirements for current and future Covid-19 patients. At present return to work efforts are under way which should have an effect on the number of cases trend. Therefore we limit our prediction to six weeks in the future. Comparing two adjacent counties may generate future efforts in equalizing the disease outcome.

## RESULTS AND DISCUSSION

Covid-19 case data for Santa Clara and San Mateo counties were obtained from the county websites [3,4]. The numbers of regular and ICU hospital beds were obtained from the California Health and Human Services websites [5]. The hospital data is available from April 1^st^, 2020.

Awareness for the mechanism of Covid-19 disease spreading through physical contact was present in the Silicon Valley since February 2020 due to many business connections with the Asian countries where the first outbreak occurred and due to the Covid-19 case reports from the Washington state. Local businesses started social distancing measures since early March at varying degrees. Shelter in place orders were issued to be in effect from March 17^th^ at six San Francisco bay area counties including the two counties in study.

The first cases of Covid-19 were reported on January 26^th^ and March 4^th^ in Santa Clara and San Mateo respectively. The Santa Clara county data could be fitted well with March 3^rd^ to be the first date (n_1_) of transmission rate decrease, 8 days for exponential time constant for transmission rate decrease (G) and 4.5% for the final transmission rate (T_f_). The San Mateo county data could be fitted well with March 11^th^ to be the first date (n_1_) of transmission rate decrease, 12 days for exponential time constant for transmission rate decrease (G) and 4.5% for the final transmission rate (T_f_). The comparison between the model predictions until June 30^th^ and actual data until May 15^th^ on number of cases, deaths and hospitalizations are shown in figures 1-5. The figures demonstrate that the model fits the actual data very well.

**Figure 1.**
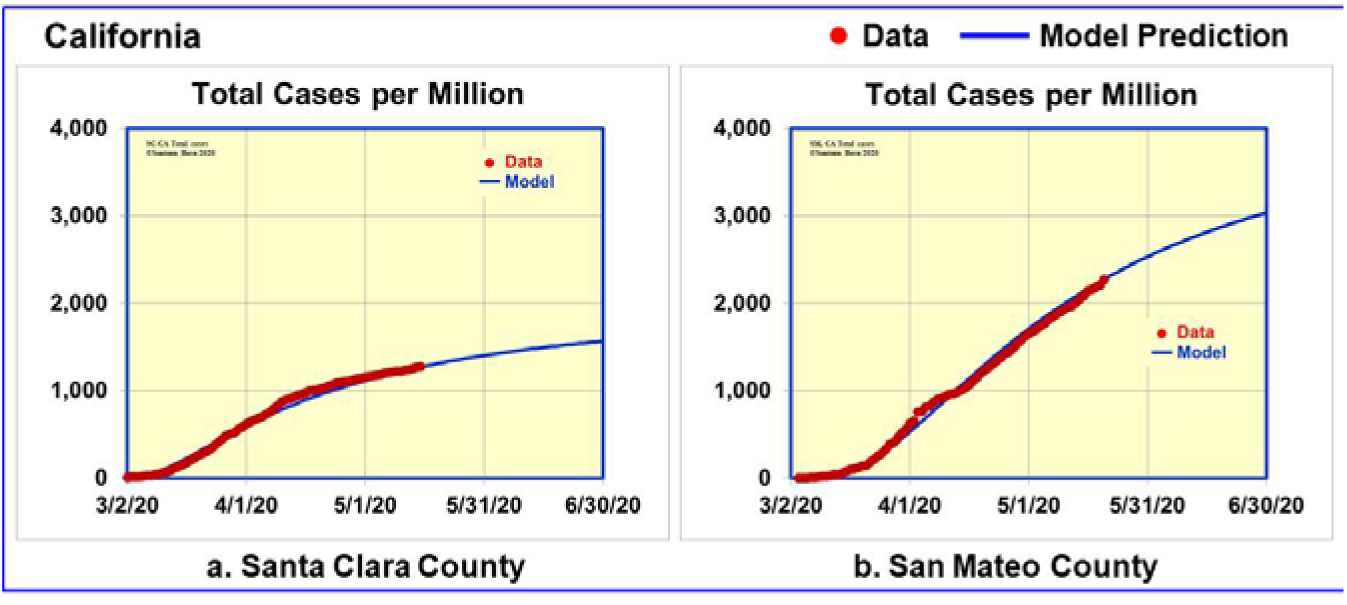
Data and model predictions for Covid-19 cases in Santa Clara and San Mateo counties

Figure 1 compares the number of Covid-19 cases per million between Santa Clara and San Mateo counties. Santa Clara is projected to have 1,500 cases per million while San Mateo is projected to have 3,000 cases per million, a factor of two more cases by June 30^th^. Santa Clara is projected to have 3,000 cases (range 2,800-3,100) while San Mateo is projected to have 2,300 cases (range 2,150-2,550). Figure 1 clearly shows that while Santa Clara case numbers have begun to plateau, San Mateo case numbers are still on the rise. Such difference in the case status can be considered by the authorities making decisions on the level and type of allowed social interactions.

Figure 2 compares the number of Covid-19 deaths per million between Santa Clara and San Mateo counties. Santa Clara is projected to have 90 deaths per million while San Mateo is projected to have 44% more deaths per million (130) by June 30^th^. The projected total number of deaths on June 30^th^ are 165 (range 155-170) and 100 (range 95-105) in Santa Clara and San Mateo respectively.

**Figure 2.**
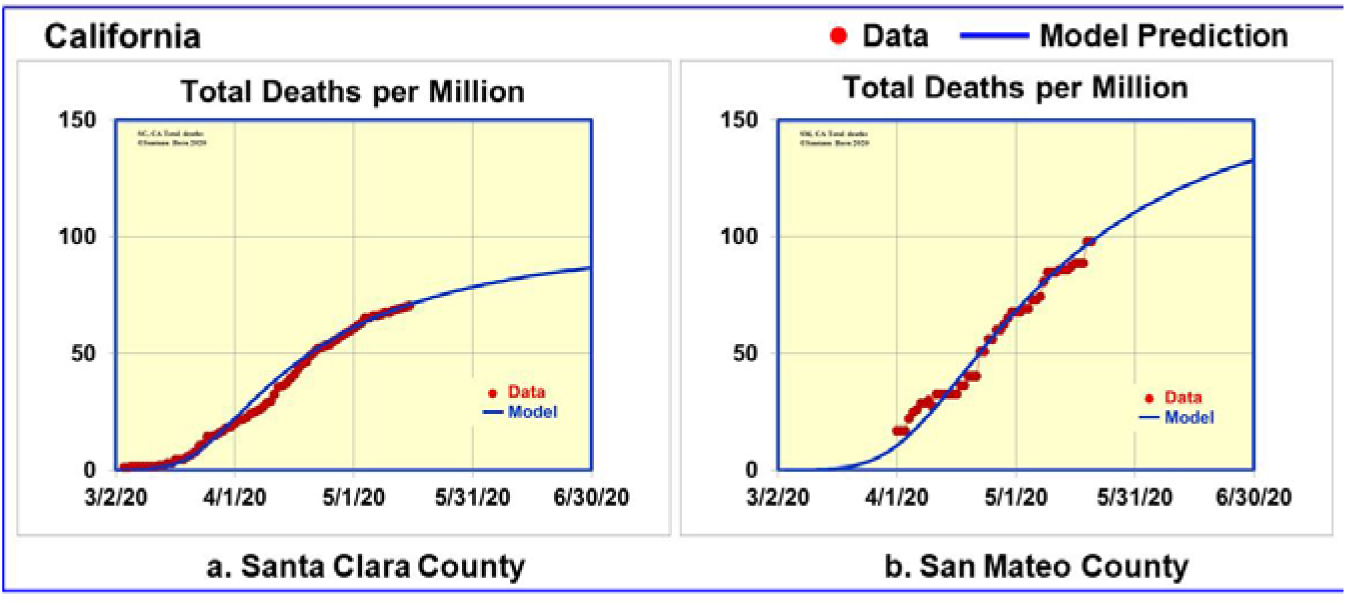
Data and model predictions for Covid-19 deaths in Santa Clara and San Mateo counties

Figures 3 and 4 compare the number of daily cases and daily deaths per million respectively between Santa Clara and San Mateo counties. San Mateo data shows spikes in daily numbers while Santa Clara numbers have less daily variation. The model predicts that in both counties, the daily cases should decrease to less than 10 (range 5-16) and daily deaths should decrease to less than 1 by June 30^th^ which will be a significant milestone.

**Figure 3.**
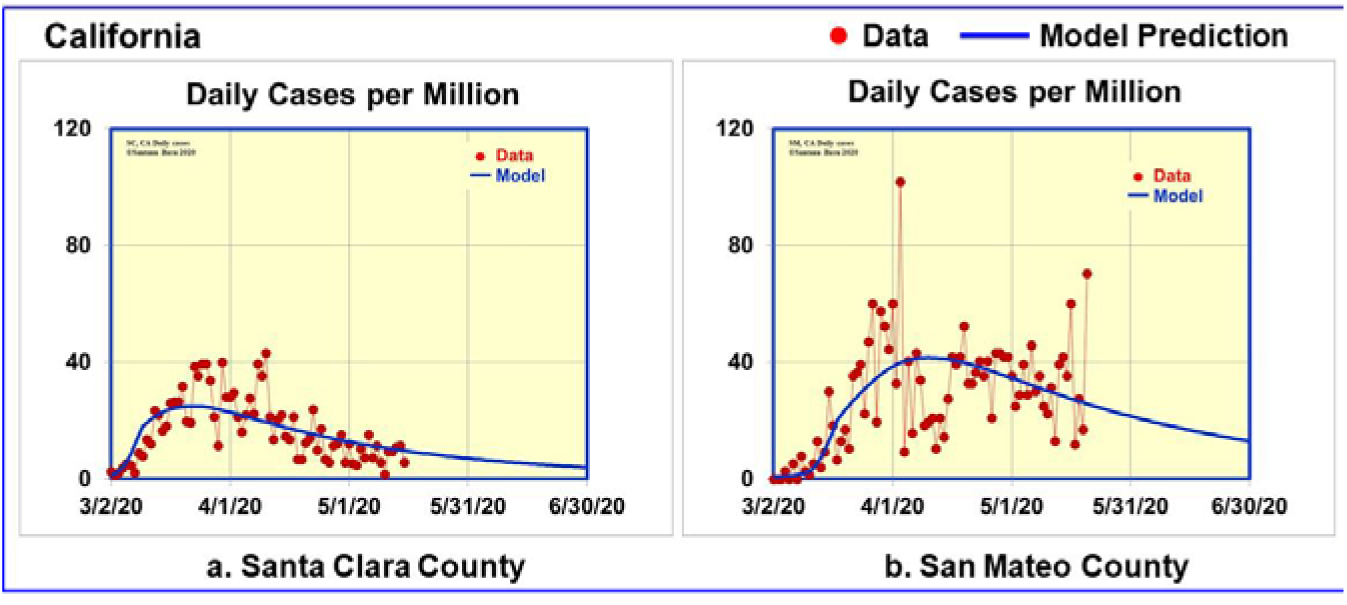
Data and model predictions for Covid-19 daily cases in Santa Clara and San Mateo counties

**Figure 4.**
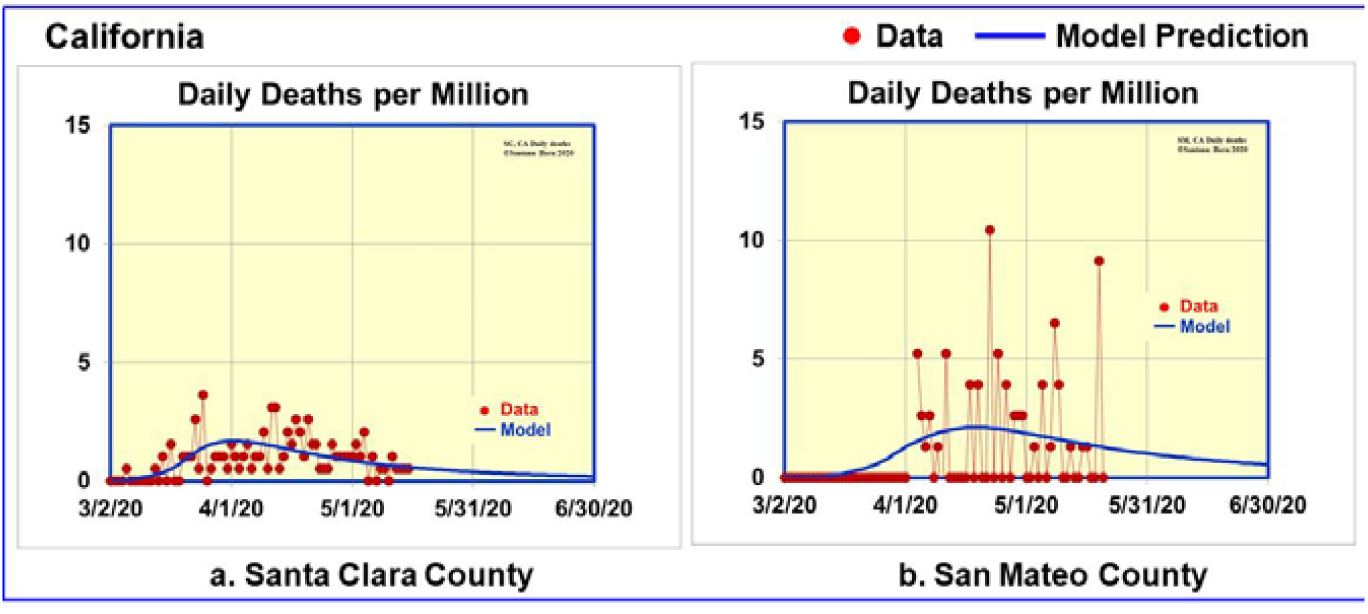
Data and model predictions for Covid-19 daily deaths in Santa Clara and San Mateo counties

Figure 5 shows the comparison between the model predictions until June 30^th^ and actual data until May 15^th^ [5] on number of hospitalizations. We analyzed the data in [5] to determine daily variation of hospitalization rate and the ratio of ICU beds to the total number of hospital beds. The model fits the data very well as shown in figure 5 by assuming 33% and 42% of the patients require ICU beds in San Mateo and in Santa Clara respectively.

**Figure 5.**
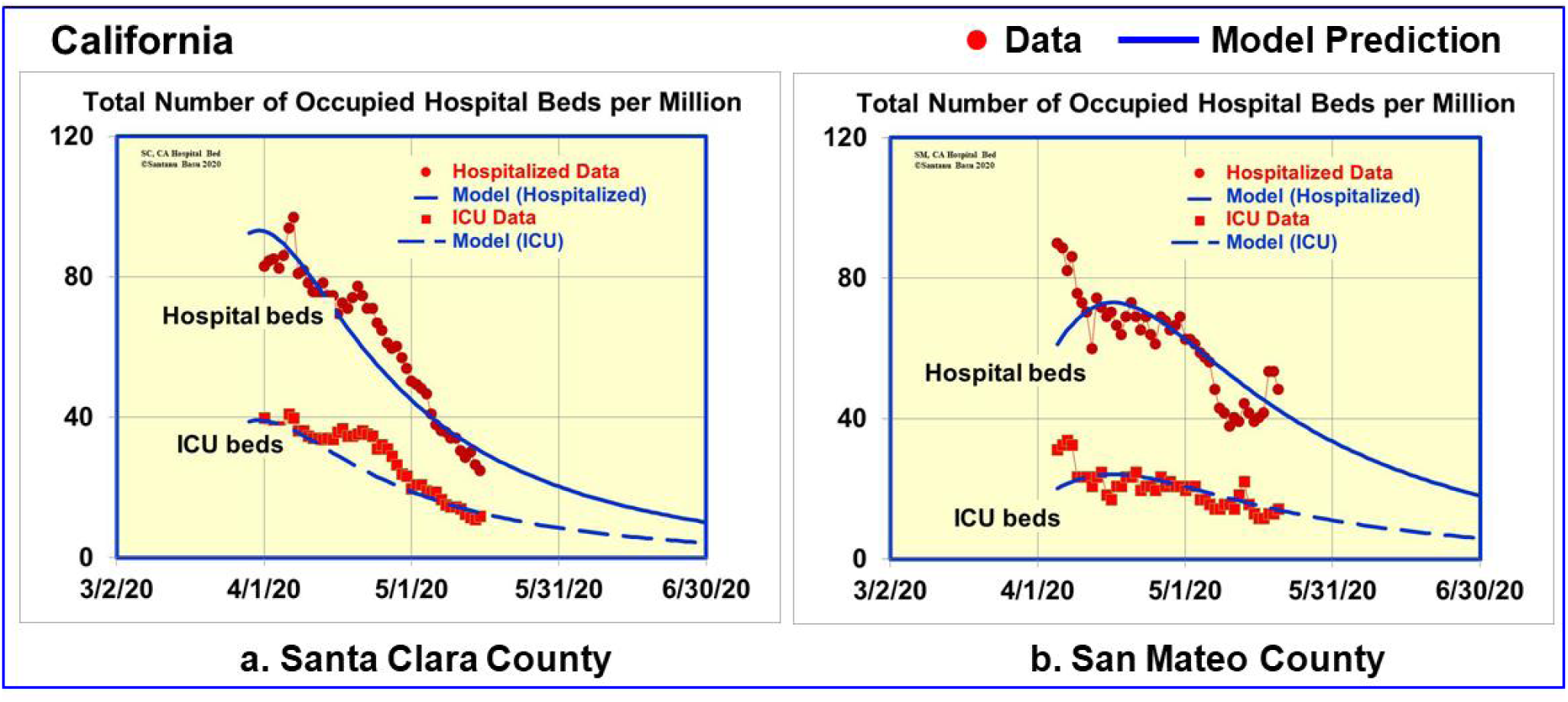
Data and model predictions for Covid-19 hospitalizations in California

For the Santa Clara county, the model predicts the number of hospitalizations to decrease from 48 on May 15^th^ to 22 (range 14-26) on June 30^th^, 2020. The number of ICU beds also will decrease from 23 on May 15^th^ to 9 (range 6-11) on June 30^th^, 2020.

For San Mateo, the model predicts the number of hospitalizations to decrease from 37 on May 20^th^ to 14 (range 9-20) on June 30^th^, 2020. The number of ICU beds also will decrease from 11 on May 20^th^ to 5 (range 3-7) on June 30^th^, 2020.

It is to be noted that the model does not take into account anticipated reopening of businesses in California around the last week of May. It will most likely increase the transmission rate parameter in the model that will be reflected in the trend in the number of cases. The model however enables one to make what if exercises such as the effect of relaxing of social distancing.

## DISCUSSION

It is very interesting that the same new virus without a vaccination caused different amount of casualties in different countries, in different states in the same country and in different regions in the same state. A part of the casualty is related to the insufficient knowledge about the virus and the transmissibility and lethality of the virus. However a significant part (by more than one or two orders of magnitude) of how many humans are infected with the virus is due to the health measures taken by the authorities and the local population. In retrospect it will be imperative to learn from the data and various modelling exercises to take the necessary steps to minimize human casualties.

Table I shows the comparison between the Santa Clara county and the San Mateo county in estimated casualties from Covid-19 by June 30^th^, 2020. New Zealand with a similar socio-economic structure as the Silicon Valley has been a model of success in minimizing human suffering from Covid-19. The data for New Zealand is also shown in the same table for comparison. If we define a factor f_NZ_, which is 1 for New Zealand, then

f_NZ_ (total cases per million) = 5 for Santa Clara and 10 for San Mateo, and

f_NZ_ (total deaths per million) = 20 for Santa Clara and 30 for San Mateo

The goal for all regions should be to achieve f_NZ_ = 1 for the next viral outbreak.

**Table 1.** Snapshot comparison of Santa Clara county, San Mateo county and New Zealand

|  |  | Santa Clara County | San Mateo County | New Zealand |
| --- | --- | --- | --- | --- |
| Population | Million | 1.93 | 0.77 | 4.78 |
| Population density | per sq mi | 1,478 | 1,030 | 46 |
| Total Cases (projected) | on 6-30-2020 | 2,976 | 2,328 |  |
| Total Deaths (projected) | on 6-30-2020 | 166 | 102 |  |
| Daily Cases (projected) | on 6-30-2020 | 8 | 10 |  |
| Daily Deaths (projected) | on 6-30-2020 | 0.4 | 0.4 |  |
| Total hospital beds (projected) | on 6-30-2020 | 22 | 14 |  |
| ICU beds (projected) | on 6-30-2020 | 9 | 5 |  |
| Cases per million (projected) | on 6-30-2020 | 1,542 | 3,023 | 314 |
| Deaths per million (projected) | on 6-30-2020 | 86 | 132 | 4 |

In conclusion, we have developed a mathematical model that provides insight into the Covid-19 cases at the county level. The model provides valuable information on hospitalization needs in the near future. Since conditions change daily which causes the model parameters to change, we do not predict more than six weeks in advance. Overall the model makes the hopeful conclusion that Covid-19 can be effectively contained by following successful examples.

The author wishes to acknowledge valuable suggestion from Professor Nigam Shah of Stanford University to look into the hospitalization needs.

## Data Availability

The data is available in the paper and the three referenced websites (linked below).

https://www.sccgov.org/sites/covid19/Pages/dashboard.aspx#cases

https://www.smchealth.org/post/san-mateo-county-covid-19-data-1

https://data.chhs.ca.gov/dataset/california-covid-19-hospital-data-and-case-statistics/resource/6cd8d424-dfaa-4bdd-9410-a3d656e1176e

## Additional Information

Correspondence and requests for materials should be addressed to the author.

